# Climatic factors influence COVID-19 outbreak as revealed by worldwide mortality

**DOI:** 10.1101/2020.04.20.20072934

**Authors:** Claudio S. Quilodrán, Mathias Currat, Juan I. Montoya-Burgos

**Affiliations:** Department of Zoology, University of Oxford, Oxford, United Kingdom; Laboratory of anthropology, genetics and peopling history, Department of Genetics and Evolution - Anthropology Unit, University of Geneva, Geneva, Switzerland; Laboratory of vertebrate evolution, Department of Genetics and Evolution, University of Geneva, Geneva, Switzerland; Institute of Genetics and Genomics in Geneva (IGE3), Switzerland

## Abstract

The COVID-19 outbreak is triggering a global crisis that is challenging governments, health systems and the scientific community worldwide^1^. A central question in the COVID-19 pandemic is whether climatic factors modulate its progression. This information is key to epidemiologists and healthcare decision-makers for improving their management plans^2^. Previous attempts to assess the impact of climatic parameters have yielded ambiguous results, either because they were using geographically or temporally restricted data, or because they were comparing infection rates across countries, which were measured differently depending on local screening strategies. In March 2020, the spread of COVID-19 dramatically increased the number of countries recording deaths^3^, providing an opportunity to use mortality rate, which is measured more homogeneously across countries than infection rates, as a descriptor of the COVID-19 outbreak over a large latitudinal range. Here, using data recorded in 208 territories from 88 countries, we show that mortality rate is negatively influenced by warmer air temperature and positively affected by higher relative humidity. Each additional Celsius degree decreases mortality rate by ∼4%, while a 1% increase in relative humidity raises mortality rate by ∼2%. Temperature is positively correlated with UV-index, for which one unit of increase results in a ∼15% decrease in the mortality rate. We also show that other factors contribute to the dynamics of the COVID-19 outbreak, such as the proportion of vulnerable age classes in the population, access to a non-overwhelmed health system, as well as governmental travel restrictions for controlling the spread of the disease. All of them are critical factors impacting the mortality rate of COVID-19. The influence of climatic factors is a warning to all southern hemisphere countries where winter is coming soon. Northern hemisphere countries should also be warned that climatic factors alone will likely not be sufficient to contain the disease.

## Introduction

The current COVID-19 outbreak is due to the coronavirus SARS-CoV-2 that originated in the area of Wuhan, Hubei province, in China^4,5^. On March 11, COVID-19 was officially declared a pandemic by the World Health Organization (WHO), as it spread extremely rapidly since December 2019, following an exponential curve worldwide^6^. As of April 20, the pandemic provoked more than 2.4 million diagnosed infections worldwide, and at least 170,000 deaths^3^. The health, social and economic consequences of this pandemic are huge^7^, with more than three billion humans under different regimes of containment at the beginning of April 2020. If most of the infections occur in the northern hemisphere, the southern hemisphere is not spared by the disease, despite the summer season. Indeed, many viral respiratory infections show seasonal fluctuation, especially in temperate regions^8,9^. Air temperature and humidity have been shown to influence the spread of influenza viruses^10^, as well as the coronaviruses SARS-CoV^11^, responsible of the severe acute respiratory syndrome (SARS) and MERS*-*CoV^12^, responsible of the Middle East respiratory syndrome.

One debated question about COVID-19 is whether its spread is being affected by season-related climatic factors, as are other seasonal viruses^2^. This is of utmost importance, as the possible climatic dependency of COVID-19 could help decision-makers to adopt the most suitable strategy to control the disease in the near future. There are several recent studies that have tackled this question since the onset of the COVID-19 outbreak, available on online archive repositories. As for other viruses, temperature and humidity seem to be factors with some influence on the spread of SARS-CoV-2, but recent studies led to contrasting results. In general, a negative relationship is found between air temperature and the spread of SARS-CoV-2 virus^13,14^, but a positive one^15^ and a lack of effect^16^ have also been reported. In the case of humidity, limited^17,18^, positive^19^ and negative^13^ effects on the infection rate have been reported. Almost all of those studies were restricted to data from China at the early stage of the epidemic. The few exceptions^13,18,19^ considered up to 47 countries, but the time frame was limited at a maximum to February the 29^th^ and these studies were based on the number of COVID-19 confirmed positive cases, which is unevenly assessed among countries.

To analyse the potential effects of climatic factors on the COVID-19 pandemic, an accurate descriptor of the outbreak that can be compared among countries is needed. The main descriptors that have been used to date include the cumulative number of reported COVID-19 positive cases at a given date^20^, the daily number of new positive cases^14,21,22^, and the daily mean number of positive cases for a given time period^19^. Yet, these descriptors are all based on the number of COVID-19 positive patients, which in turn depends on the countries’ screening strategies.

As of 20 March, many countries were testing patients exhibiting clear COVID-19 symptoms, such as Italy, Spain, France, Belgium, Switzerland, the UK, and the USA. Other countries, such as South Korea, Singapore, Australia, United Arab Emirates and more recently Germany, have opted to screening their population more widely. It results that countries performing more tests are either following a strategy of wide screening, whatever the proportion of positive patients, or are having numerous patients with clear COVID-19 symptoms. These two contrasting situations lead to infection estimates that are hardly comparable among countries.

To show that descriptors based on COVID-19 positive patients might be strongly biased when analysing data reported from multiple countries, we performed a correlation analysis between the mean maximum air temperature between mid-February and mid-March 2020 and the total number of tests performed per country, as of March 20. We found a strong negative correlation, where countries with lower temperatures tend to perform more tests (*r* =-0.48, n = 41, *p* value = 0.001). This correlation might be influenced by the higher impact of this disease in the northern hemisphere during the first months of 2020. However, the reasons for this correlation remain unclear, and may lead to temperature-biased numbers of COVID-19 positive patients across countries. Therefore, we argue that descriptors based on the number of tests performed should not be used to assess the contribution of air temperature (or other climatic factors) in the trans-country COVID-19 outbreak.

To reach a better comprehension of the COVID-19 pandemic, the global extent of the disease in March 2020 brings new data to test the effects of climatic factors. Indeed, the number of countries recording deaths increased from nine by the end of February 2020 to more than 120 by the end of March 2020^3^, providing a first-hand opportunity to use the mortality rate as the descriptor of COVID-19 outbreak over a large latitudinal range. Here we use the mortality rate expressed as the number of deaths per inhabitant over a standardized time period of 15 days, corresponding to the 15 days following the first reported COVID-19 death in the country. This metric is much less dependent on the local testing strategy. In almost all countries, the number of COVID-19 deaths publicly reported corresponds to the cases of COVID-19 positive patients and patients displaying clear diagnostic COVID-19 symptoms who died on a given date. Although this number may not reflect the exact number of deaths due exclusively to COVID-19 infection, because death can result from other underlying health problem exacerbated by the viral infection or because patients may display symptoms that are mistakenly attributed to COVID-19 (ascertainment bias), these biases are similar across countries allowing credible country-wise comparisons. Consequently, mortality rate is a reliable descriptor for the worldwide COVID-19 outbreak. However, when it comes to use the mortality rate as the outbreak descriptor, it implies that the disease is well established, and thus we included other known explanatory factors in our analyses: the proportion of the most susceptible age group in the population, the number of beds in hospitals (reflecting access to health care), and the timing and strength of governmental travel measures for controlling the spread of the disease.

## Results

We examined the possible effects of three climatic variables on the worldwide COVID-19 outbreak: air temperature, UV-index and relative humidity. All these variables are averaged over 30 days, which are the 15 days before and the 15 days after the first registered death in each territory or country. We used a linear mixed model (LMM) with the log transformed mortality rate per region as the response variable. The model considers the country-level as a random intercept and implements an exponential geographical correlation structure (see Methods). We included the climatic variables together with other variables that may explain the extent of the epidemic outbreak, namely (i) the proportion of the population older than 64 years old; (ii) the number of beds in hospitals (per 10K inhabitants), and (iii) the timing and strength of the governmental travel measures to control the spread of the disease. As air temperature is highly correlated with UV-index (see Methods), we tested a temperature-based model (excluding UV-index) and a UV-based model (excluding temperature). We ranked candidate models that can be built with combinations of the six variables according to their resulting weighted AIC (*ω*_*AIC*_)^23^. We then computed a model average of all candidate models included in a confidence set of 95% (Σ*ω*_*i*_ ≥ 0.95) (Table S1, Supporting Information).

Very similar results were obtained with the temperature-based and the UV-based averaged models, yet with slightly higher significance values for the latter (Table 1). Both approaches resulted in sets of best models explaining between 12% and 25% of variation in mortality rate by the fixed variables (R^2^ _GLMM(m)_, Table S1), and between 49% and 54% when considering both fixed and random variables (R^2^ _GLMM(c)_, Table S1). In these models, temperature and UV-index influenced negatively the mortality rate, while the opposite was observed for the relative humidity (Fig. 1). All other explanatory variables also contributed to explain the COVID-19 outbreak. The proportion of the population older than 64 years old had a positive effect on mortality rate, while the number of beds in hospitals and the travel restrictions implemented in each country showed a negative effect (Table 1).

**Table 1.** Results of the model averaging procedure based on the best candidate models. The models show the effect of the descriptor variables on the mortality rate during the COVID-19 outbreak. The results for the temperature-based average model and the UV-based average model are presented (see Methods).

| Model | Variable | Estimate | SE | DF | *t* | *p*-value |
| --- | --- | --- | --- | --- | --- | --- |
| Temperature | Intercept | -15.56 | 1.09 | 1.09 | 14.22 | <0.01 |
|  | Temperature | -0.04 | 0.02 | 0.02 | 2.47 | 0.01 |
|  | Humidity | 1.70 | 0.86 | 0.87 | 1.95 | 0.05 |
|  | Above 64 years old | 0.11 | 0.04 | 0.04 | 2.69 | 0.01 |
|  | Beds in hospitals | -0.02 | 0.01 | 0.01 | 2.09 | 0.04 |
|  | Restrictions:middle | 0.07 | 0.44 | 0.44 | 0.15 | 0.88 |
|  | Restrictions:strong | -0.92 | 0.47 | 0.47 | 1.94 | 0.05 |
| UV index | Intercept | -15.17 | 1.29 | 1.29 | 11.73 | <0.01 |
|  | UV index | -0.16 | 0.06 | 0.06 | 2.60 | 0.01 |
|  | Humidity | 1.85 | 0.87 | 0.87 | 2.12 | 0.03 |
|  | Above 64 years old | 0.11 | 0.04 | 0.04 | 2.66 | 0.01 |
|  | Beds in hospitals | -0.02 | 0.01 | 0.01 | 2.13 | 0.03 |
|  | Restrictions:middle | 0.10 | 0.43 | 0.44 | 0.22 | 0.82 |
|  | Restrictions:strong | -0.96 | 0.46 | 0.47 | 2.04 | 0.04 |

**Figure 1.**
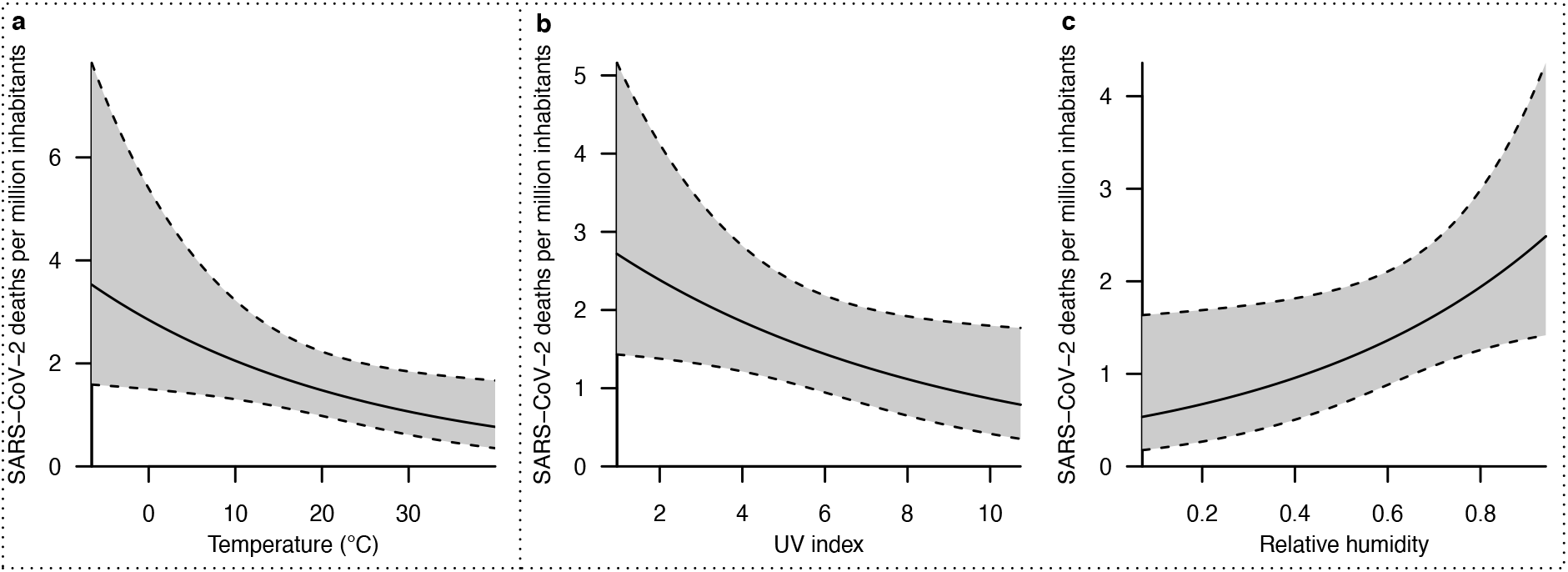
Climatic factors influencing the mortality rate during the COVID-19 outbreak (calculated per million inhabitants). A) effect of temperature (in degrees Celsius), B) effect of UV-index, C) effect of relative humidity. Because temperature and UV-index are highly correlated (see methods), these two variables were analysed separately in two distinct models. The influence of humidity is taken from the UV-based model.

The results for the temperature-based averaged model showed that every extra degree Celsius decreases the mortality rate by 3.74% (*e*^-0.04^-1, Table 1). The UV-based averaged model showed that every unit increase in UV-index results in a 14.72% decrease in the mortality rate (*e*^-0.16^-1, Table 1). In this last model, humidity had a positive effect, in which every 1% increase of relative humidity results in a 1.85% mortality rate increase (*e*^-1.85·0.01^-1, Table 1). Regarding the other variables in our models, every 1% increase in the proportion of the population older than 64 years old increased the mortality rate by 11.31% (*e*^0.11^-1, Table 1). Every extra bed in hospitals for 10K inhabitants decreased the mortality rate by 2.22% (*e*^-0.02^-1, Table 1). Finally, strong and early governmental travel restrictions resulted in a geometric mean of mortality rate 61.8% lower than when restrictions were weak and imposed late (*e*^-0.96^-1, Table 1). No effect is observed concerning intermediate timing and strength of governmental travel restrictions (Table 1).

## Discussion

We stress that warm air temperatures, intense UV light, or low relative humidity may act by reducing the transmission of the SARS-CoV-2 virus or it may directly or indirectly inactivate this virus. Preliminary results indicate that, under laboratory conditions, the SARS-Cov-2 virus survives less than 4 days at 37° C^24^ while at 4 °C it may remain stable for more than 14 days^25^, offering one possible explanation to the negative relation between air temperature and the COVID-19 mortality rate. Some evidence suggests that the transmission efficiency of influenza viruses by respiratory droplet increases when air is cold and dry^26^. Moreover, in periods of the year or places of the world with cold and dry air and reduced UV light, other human respiratory diseases are more frequent^27^ and may facilitate inter-individual transmission via coughing or sneezing. When the weather is cold, people also tend to spend more time in closed and populated environments, which also facilitates inter-individual transmission. A causative link between exposure to sunlight (including UV light), a higher synthesis of vitamin D, and better efficiency of the human immune system has been evidenced^28^, which may explain in part the negative relation between the UV-index and the COVID-19 mortality rate. Yet, these modes of action remain hypotheses to be tested in the case of COVID-19.

Humankind is facing an unprecedented worldwide crisis, forcing decision makers to move forward into the labyrinth of unknowns. Understanding how climatic factors have contributed to the COVID-19 outbreak until now could strengthen the projections of its future spread. Previous analyses were unable to provide an unambiguous view because of limited data or biased descriptors for country-wise comparisons. Using global datasets and a homogeneous descriptor of the COVID-19 outbreak, we show that air temperature and UV-index are related climatic variables that play a substantial role in explaining the COVID-19 mortality rate. A higher value in one or other of the variables is associated with lower mortality rates. Relative humidity is a third climatic variable able to explain a reduction in COVID-19 mortality rate when it is lower. We also show the importance of age class structure, access to healthcare systems and governmental travel restrictions in explaining the COVID-19 outbreak. Although our results may somewhat relieve some of the anxiety of people living in regions entering the warm season or living in warm countries, it is clear that climatic factors alone will not passively stop the outbreak. However, our results do represent a warning for the populations and decision-makers of countries with cold climates or moving towards the cold season. Importantly, the clear contribution of climatic factors on the COVID-19 outbreak, however, does not ensure that this disease will evolve into a winter-seasonal epidemic viral disease like the influenza virus^29^. More research is urgently needed to better understand the dynamics of the COVID-19 pandemic in order to save lives.

## Methods

### COVID-19 dataset

We collected the number of deaths per day due to the COVID-19 in each country affected by SARS-CoV-2 from the dataset hosted by the Centre for Systems Science and Engineering at Johns Hopkins University^3^. This dataset is maintained daily and is verified by registers provided by various health state authorities as well as the World Health Organisation (WHO). The dataset provides information at the country level but also about sub-regions of China, The United States, Canada, and Australia. French and British overseas territories are also included separately in this dataset. Due to the uncertainly regarding the initial stage of the spread of this disease^4^, we decided to exclude the dataset for mainland China. We considered the date of first registered death in each country as well as the cumulative number of deaths during the first 15 days of deaths registration in each country or territory. We used the cumulative mortality at day 15 in order to include the maximum available information and to compare data extracted from similar epidemic phases across countries. We considered all regions registering at least 15 days of deaths until 6^th^ April 2020. The final dataset includes 208 territories in 88 countries.

### Climatic dataset

The historical weather conditions in each country or territory included in the COVID-19 dataset were obtained from the Dark Sky API dataset by using the R package “darksky”^30^. We considered the geographical coordinates associated with each record in the COVID-19 dataset. We included in the analysis the maximum temperature (°C), relative humidity, and UV-index. These variables were taken by day for the 15 days before and the 15 days after the first registered death in each country or territory recorded. We computed the average across these 30 days for each climatic variable.

### Non-climatic dataset

The proportion of the population older than 64 years old per country was calculated on the basis of the population indicators available from the World Bank database^31^. The indicator of the number of hospital beds per 10K inhabitants was obtained from the Global Health Observatory (GHO) database of the World Health Organization (WHO)^32^. We used the latest available data per country, which varied from 2005 to 2015, but most data were from the period 2010-2015. Governmental immigration restrictions per country were used as a proxy for the governmental travel measures for controlling the spread of the disease. This last dataset was taken from the COVID-19 Travel Restriction Monitoring database, compiled by the International Organization for Migration (IOM)^33^. We added some missing data directly from the primary source database, the International Air Transport Association (IATA)^34^. For each country and date, from March 8 to March 26, we summed the number of other countries for which immigration restrictions were imposed. We considered all the IOM traveling restriction categories with a link to COVID-19 spread control. Based on these daily immigration restrictions, we grouped the countries into three categories. Category 1 (soft restriction) includes countries that imposed virtually no restrictions or otherwise late in time, not before March 18, and only on a subset of countries. Category 2 (middle restriction) contains the countries that started imposing noticeable immigration restrictions from March 11 to 17 with a fast daily-increase in the number of countries with restricted entrance. Category 3 (strong restriction) groups the countries that decided immigration restrictions early in time, before March 11, with a fast daily-increase in the number of countries from which immigration was restricted. The rates of COVID-19 tests performed per country were obtained from the database of STATISTA^35^.

### Statistical analysis

Our response variable (mortality rate) is the number of deaths per inhabitant during the first 15 days of deaths registration in each country or territory included in the COVID-19 dataset. This variable is computed by considering the cumulative number of deaths on day 15 divided by the population size of each country or region. This variable was log-transformed in order to maintain a linear relationship with the predictor variables. The fixed predictor variables included temperature, UV-index, relative humidity, proportion of the population older than 64 years old, number of beds in hospitals for every 10K inhabitants, and the governmental travel measures implemented in each country to contain the spread of this disease. Since temperature and UV-index are highly correlated (*r* ∼ 0.9, *p-value* < 0.001), we included those variables in two separated models. We verified the collinearity among predictor variables by computing the variance inflator factor (VIF). All variables included in the separate models with either temperature or UV-index did not show collinearity with other predictor variables (VIFs < 1.6)^36^. We also estimated the effect of including a spatial correlation structure as well as the effect of the country-level as a random effect, following Zuur, et al. ^36^. The model selection was based on the lowest Akaike Information Criterion value (AIC)^23^. The final model structure was a linear mixed model (LMM) that considers an exponential spatial correlation and country-level as a random intercept. We explored all combinations of predictor fixed variables. These models were ranked by using the Akaike weights (*ω*_*i*_), which provide a measure of the probability of each model being the most suitable among all of the candidate models. Since there is no single model that has a high Akaike weight (*ω*_*i*_ > 0.95), we averaged all models in a confidence set at 95% (Σ*ω*_*i*_ ≥ 0.95)^23^. We reported the conditional (*R*^*2*^_*GLMM(c)*_) and marginal (*R*^*2*^_*GLMM(m)*_) coefficient of determination for each model included in the set of candidate best models included in the model averaging. These values represent the variance explained by the fixed predictors and both the fixed predictors and the random variable, respectively^37^. In the case of the relationship between the number of COVID-19 tests and the temperature of each country, the model with the lowest AIC was a simple linear relationship without the influence of a spatial correlation structure or the inclusion of a country-level random effect. All statistical analyses were performed using *R* 3.6.2^*38*^. We used the package ‘nlme’^39^ for the linear mixed models and the package ‘MuMIn’^40^ for the model averaging and the computation of the determination coefficients.

## Data Availability

The datasets generated and analysed in the current study are available from the corresponding author upon request

## Acknowledgements

All authors are supported by grants from the Swiss National Science Foundation: n° 31003A_182577 to MC, n° P400PB_183930 to CSQ, and 310030_185327/1 to JMB.

## Author contributions

C.S.Q., M.C. and J.I.M.B. conceived the original idea. C.S.Q. performed the statistical analysis. All authors analysed and discussed the results, and contributed critically in the writing of the manuscript.

## Supporting Information

**Table S1.** Fixed variable selection for explaining the mortality rate during the COVID-19 outbreak. The best models, according to the weighted AIC criterion, for the analysis including the air temperature and the analysis including the UV-index, are reported. These models are considered in the model averaging (see Table 1). For the model averaging, we considered all models with a cumulative weighted AIC of 0.95 (Σ*ω*_AIC_ ≥ 0.95).

| Model | Fixed variable* | df | AICc | $\Delta AICc$ | $\omega_{AIC}$ | $R^2_{GLMM(m)}$ | $R^2_{GLMM(c)}$ |
| --- | --- | --- | --- | --- | --- | --- | --- |
| Temperature | beds+hum+65old+temp | 8 | 720.17 | 0.00 | 0.29 | 0.21 | 0.51 |
|  | beds+65old+temp | 7 | 721.28 | 1.11 | 0.17 | 0.19 | 0.52 |
|  | beds+hum+65old+rest+temp | 10 | 721.39 | 1.22 | 0.16 | 0.25 | 0.52 |
|  | beds+65old+rest+temp | 9 | 722.17 | 2.00 | 0.11 | 0.24 | 0.53 |
|  | beds+hum+65old | 7 | 722.98 | 2.81 | 0.07 | 0.18 | 0.52 |
|  | hum+65old+rest+temp | 9 | 724.02 | 3.85 | 0.04 | 0.19 | 0.52 |
|  | beds+hum+65old+rest | 9 | 724.11 | 3.93 | 0.04 | 0.22 | 0.52 |
|  | 65old+rest+temp | 8 | 725.01 | 4.84 | 0.03 | 0.17 | 0.53 |
|  | hum+65old+temp | 7 | 725.02 | 4.85 | 0.03 | 0.14 | 0.52 |
|  | beds+65old | 6 | 725.08 | 4.90 | 0.02 | 0.16 | 0.54 |
|  | hum+65old+rest | 8 | 725.40 | 5.23 | 0.02 | 0.17 | 0.52 |
|  | beds+65old+rest | 8 | 725.87 | 5.70 | 0.02 | 0.20 | 0.54 |
|  | hum+rest+temp | 8 | 726.30 | 6.13 | 0.01 | 0.15 | 0.50 |
| UV index | beds+hum+65old+uv | 8 | 720.99 | 0.00 | 0.28 | 0.20 | 0.51 |
|  | beds+hum+65old+rest+uv | 10 | 721.82 | 0.83 | 0.18 | 0.25 | 0.51 |
|  | beds+hum+65old | 7 | 722.98 | 1.99 | 0.10 | 0.18 | 0.52 |
|  | beds+65old+uv | 7 | 723.28 | 2.29 | 0.09 | 0.18 | 0.52 |
|  | beds+65old+rest+uv | 9 | 723.75 | 2.76 | 0.07 | 0.23 | 0.52 |
|  | beds+hum+65old+rest | 9 | 724.11 | 3.12 | 0.06 | 0.22 | 0.52 |
|  | hum+65old+rest+uv | 9 | 724.57 | 3.58 | 0.05 | 0.18 | 0.51 |
|  | beds+65old | 6 | 725.08 | 4.09 | 0.04 | 0.16 | 0.54 |
|  | hum+65old+rest | 8 | 725.40 | 4.41 | 0.03 | 0.17 | 0.52 |
|  | hum+rest+uv | 8 | 725.68 | 4.70 | 0.03 | 0.16 | 0.49 |
|  | beds+65old+rest | 8 | 725.87 | 4.89 | 0.02 | 0.20 | 0.54 |
|  | hum+65old+uv | 7 | 725.97 | 4.98 | 0.02 | 0.13 | 0.51 |
|  | hum+65old | 6 | 726.32 | 5.33 | 0.02 | 0.12 | 0.52 |
|  | 65old+rest+uv | 8 | 726.51 | 5.52 | 0.02 | 0.17 | 0.52 |

\*Code of each variable: air temperature (temp), relative humidity (hum), UV-index (uv), proportion of the population older than 64 years old (65old), number of beds in hospitals per 10K inhabitants (beds), country travel restrictions (rest).

## References

1 Wang, C., Horby, P. W., Hayden, F. G. & Gao, G. F. A novel coronavirus outbreak of global health concern. The Lancet 395, 470–473, doi:10.1016/S0140-6736(20)30185-9 (2020).

2 Neher, R. A., Dyrdak, R., Druelle, V., Hodcrof, E. B. & Albert, J. Potential impact of seasonal forcing on a SARS-CoV-2 pandemic. Swiss Med Wkly 150:w20224, doi:10.4414/smw.2020.20224 (2020).

3 Dong, E., Du, H. & Gardner, L. An interactive web-based dashboard to track COVID-19 in real time. Lancet Infect Dis, doi:10.1016/S1473-3099(20)30120-1 (2020).

4 Wu, F. et al. A new coronavirus associated with human respiratory disease in China. Nature 579, 265–269, doi:10.1038/s41586-020-2008-3 (2020).

5 Zhou, P. et al. A pneumonia outbreak associated with a new coronavirus of probable bat origin. Nature 579, 270–273, doi:10.1038/s41586-020-2012-7 (2020).

6 World Health Organization. Coronavirus disease 2019 (COVID-19), Situation Report -51. (World Health Organization, accessible at https://www.who.int/docs/default-source/coronaviruse/situation-reports/20200311-sitrep-51-covid-19.pdf?sfvrsn=1ba62e57_10, Geneva, 2020).

7 Cohen, J. & Kupferschmidt, K. Countries test tactics in ‘war’ against COVID-19. Science 367, 1287–1288, doi:10.1126/science.367.6484.1287 (2020).

8 Dowell, S. F. & Ho, M. S. Seasonality of infectious diseases and severe acute respiratory syndrome-what we don’t know can hurt us. Lancet Infect Dis 4, 704–708, doi:10.1016/S1473-3099(04)01177-6 (2004).

9 Li, Y. et al. Global patterns in monthly activity of influenza virus, respiratory syncytial virus, parainfluenza virus, and metapneumovirus: a systematic analysis. Lancet Glob Health 7, e1031–e1045, doi:10.1016/S2214-109X(19)30264-5 (2019).

10 Lowen, A. C., Mubareka, S., Steel, J. & Palese, P. Influenza virus transmission is dependent on relative humidity and temperature. PLoS Pathog 3, 1470–1476, doi:10.1371/journal.ppat.0030151 (2007).

11 Lin, K., Yee-Tak Fong, D., Zhu, B. & Karlberg, J. Environmental factors on the SARS epidemic: air temperature, passage of time and multiplicative effect of hospital infection. Epidemiol Infect 134, 223–230, doi:10.1017/S0950268805005054 (2006).

12 Gardner, E. G. et al. A case-crossover analysis of the impact of weather on primary cases of Middle East respiratory syndrome. BMC Infect Dis 19, 113, doi:10.1186/s12879-019-3729-5 (2019).

13 Bannister-Tyrrell, M., Meyer, A., Faverjon, C. & Cameron, A. Preliminary evidence that higher temperatures are associated with lower incidence of COVID-19, for cases reported globally up to 29th February 2020. medRxiv, doi:10.1101/2020.03.18.20036731 (2020).

14 Zhu, Y. & Xie, J. Association between ambient temperature and COVID-19 infection in 122 cities from China. Science of The Total Environment, doi:10.1016/j.scitotenv.2020.138201 (in press).

15 Ma, Y. et al. Effects of temperature variation and humidity on the mortality of COVID-19 in Wuhan. Science of the Total Environment, doi:10.1101/2020.03.15.20036426 (in press).

16 Pawar, S., Stanam, A., Chaudhari, M. & Rayudu, D. Effects of temperature on COVID-19 transmission. medRxiv, doi:doi.org/10.1101/2020.03.29.20044461 (2020).

17 Shi, P. et al. The impact of temperature and absolute humidity on the coronavirus disease 2019 (COVID-19) outbreak - evidence from China. medRxiv, doi:10.1101/2020.03.22.20038919 (2020).

18 Luo, W. et al. The role of absolute humidity on transmission rates of the COVID-19 outbreak. medRxiv, doi: https://doi.org/10.1101/2020.02.12.20022467 (2020).

19 Wang, M. et al. Temperature significant change COVID-19 Transmission in 429 cities. medRxiv, doi:10.1101/2020.02.22.20025791 (2020).

20 Bu, J. et al. Analysis of meteorological conditions and prediction of epidemic trend of 2019-nCoV infection in 2020. medRxiv, doi:10.1101/2020.02.13.20022715 (2020).

21 Araújo, M. B. & Naimi, B. Spread of SARS-CoV-2 Coronavirus likely constrained by climate. medRxiv, doi:10.1101/2020.03.12.20034728 (2020).

22 Wang, J., Tang, K., Feng, K. & Lv, W. High Temperature and High Humidity Reduce the Transmission of COVID-19. arxiv.org, doi:10.2139/ssrn.3551767 (2020).

23 Burnham, K. P. & Anderson, D. R. A practical information-theoretic approach. Model selection and multimodel inference, 2nd ed. Springer, New York (2002).

24 World Health Organization. First data on stability and resistance of SARS coronavirus compiled by members of WHO laboratory network, <https://www.who.int/csr/sars/survival_2003_05_04/en/> (2020).

25 Chin, A. et al. Stability of SARS-CoV-2 in different environmental conditions. medRxiv (2020).

26 Lowen, A. C. & Steel, J. Roles of humidity and temperature in shaping influenza seasonality. Journal of virology 88, 7692–7695 (2014).

27 Hyrkäs-Palmu, H. et al. Cold weather increases respiratory symptoms and functional disability especially among patients with asthma and allergic rhinitis. Scientific reports 8, 1–8 (2018).

28 Aranow, C. Vitamin D and the immune system. Journal of investigative medicine 59, 881–886 (2011).

29 Tamerius, J. D. et al. Environmental predictors of seasonal influenza epidemics across temperate and tropical climates. PLoS pathogens 9 (2013).

30 Rudis, B. darksky: Tools to Work with the ‘Dark Sky’ ‘API’. R package version 1.3.0., <https://CRAN.Rprojectorg/package=darksky.> (2017).

31 The World Bank. Wold Bank Statistics: population ages 65 and above (% of total population), <https://data.worldbank.org/indicator/SP.POP.65UP.TO.ZS?view=chart> (2019).

32 World Health Organization. The Global Health Observatory: Hospital beds (per 10 000 population) <>https://www.who.int/data/gho/data/indicators/indicator-details/GHO/hospital-beds-(per-10-000-population)> (2020).

33 International Organization for Migration. Mobility Impact COVID-19, <https://migration.iom.int> (2020).

34 International Air Transport Association. Government Measures Related to Coronavirus (COVID-19), <https://www.iata.org/en/programs/safety/health/diseases/government-measures-related-to-coronavirus> (2020).

35 Statista. Rate of coronavirus (COVID-19) tests performed in select countries worldwide as of March 20, 2020 (per one million population), <https://www.statista.com/statistics/1104645/covid19-testing-rate-select-countries-worldwide/> (2020).

36 Zuur, A., Ieno, E. N., Walker, N., Saveliev, A. A. & Smith, G. M. Mixed effects models and extensions in ecology with R. (Springer Science & Business Media, 2009).

37 Nakagawa, S., Johnson, P. C. & Schielzeth, H. The coefficient of determination R 2 and intra-class correlation coefficient from generalized linear mixed-effects models revisited and expanded. Journal of the Royal Society Interface 14, 20170213 (2017).

38 R: A Language and Environment for Statistical Computing. R Foundation for Statistical Computing, Vienna, Austria (2019).

39 Pinheiro, J., Bates, D., DebRoy, S. & Sarkar, D. R Core Team (2019) nlme: linear and nonlinear mixed effects models. R package version 3.1-142, <https://CRAN.R-project.org/package=nlme> (2019).

40 Barton, K. Package ‘MuMIn’: Multi-Model Inference. R package version 1.43.15, <https://CRAN.R-projectorg/package=darksky> (2019).

